# COVID-19 Patients Analysis using SuperHeat Map and Bayesian Network to identify Comorbidities Correlations under Different Scenarios

**DOI:** 10.1101/2021.05.11.21257055

**Authors:** O. Nolasco-Jáuregui, L. A. Quezada-Téllez, E. E. Rodriguez-Torres, G. Fernández-Anaya

## Abstract

**Background:** Given the exposure risk of comorbidities in Mexican society, the new pandemic involves the highest risk for the population in the history.

**Objective:** This article presents an analysis of the COVID-19 risk from Mexico’s regions.

**Method:** The study period runs from April 12 to June 29, 2020 (220,667 patients). The method has a nature applied and according to its level of deepening in the object of study it is framed in a descriptive and explanatory analysis type. The data used here has a quantitative and semi-quantitative characteristic because they are the result of a questionnaire instrument made up of 34 fields and the virus test. The instrument is of a deliberate type. According to the manipulation of the variables, this research is a secondary type of practices, and it has a factual inference from an inductive method because it is emphasizing the concomitant variations for each region of the country.

**Results:** Region 1 and Region 4 have a higher percentage of hospitalized patients, while Region 2 has a minimum of them. The average age of non-hospitalized patients is around 40 years old, while the hospitalized patients’ age it is close to 55 years. The most sensitive comorbidities in hospitalized patients are three principal: obesity, diabetes mellitus and hypertension. The patients whose needed the mechanical respirator were in ranged from 7.45% to 10.79%.

**Conclusions:** There is a higher risk of lose their lives in the Region 1 and Region 4 territories than in the Region 2, this information was dictated by the statistical analysis..

## INTRODUCTION

The city of Wuhan in China got the world attention dues to a new disease outbreak of respiratory illness called COVID-19. In December 2019, the Hubei Province from the Huanan Seafood Wholesale Market has been widely considered the origin of this new epidemic. The World Health Organization (WHO) receives notifications about this new disease by the Chinese authorities until December 31 (1).

Coronavirus are a large family of viruses that are can cause illness in animals and humans. In human’s case, as far as it is known, several of these coronaviruses induce usual signs of the cold and in severe cases give rise to a respiratory diseases such as Middle East Respiratory Syndrome (MERS) and Severe Acute Respiratory Syndrome (SARS) (1).

On March 11th, the 118, 000 infections cases reported globally in 114 countries with 4, 291 people have lost their lives, the WHO Director declared that COVID-19 can be characterized as a pandemic (2). As consequence of this situation, on March 16 Mexico’s health ministry announced the implementation of “healthy distance” to avoid infection. Measures to stop the spread of the coronavirus, all activities non-essential presented temporarily restrictions, except in the essential activities include the provision of medical services and supplies, assurance of public safety, maintenance of fundamental economic functions and others essential categories (3).

One of the main measures established by the Mexican government was that the population to stay at home and to prevent increased risk of infection. Involved in the priority sectors to protect are adults on ageing as well as those people with chronic conditions such as cardiovascular disease, diabetes, chronic respiratory diseases, hypertension and cancer illness. The presence of certain comorbidities may represent potential risk factors and death among patients who was infected by the virus (3). It is important to mention the previous risk scenario in Mexico before the arrival of the COVID-19. In 2018, 722, 611 deaths were reported in Mexico, of which ones 56.4% were men, approximately 43.5% were women, and the rest of them 376 cases the sex was not specified. Of the total deaths, 88.4% were health-related diseases, while 11.6% were due to other causes, mainly to accidents, homicides, and suicides. The three leading cause of death for both men and women are heart disease, diabetes mellitus and malignant tumors. Homicides were fourth cause of deaths in men (4). Research aim refers to analyze the behavior of COVID-19 in Mexico from a period to 79 days (April 12 to June 29, 2020). The study project report is a statistical analysis which is based in a compared of the contagion population with the chronic diseases of the patients, and their exposure risk to the virus. The paper is structured as follows: Summary, Introduction, and Section 2nd that is a brief overview of the arrival of COVID-19 in Mexico. Section 3rd presents a statistical analysis descriptive research of the database reported by the Mexican authorities. Section 4th shows the results obtained from the statistical data, those are discussed in this section.

### Arrives COVID-19 to Mexico

COVID-19 arrived to Mexico at the end of February 2020. Thursday 27 of February 2020, it was announced at the media that a patient had been tested positive for the coronavirus. This patient goes to the National Institute of Respiratory Diseases (INER) where he mentioned having traveled to Bergamo Italy; there he had contact with an infected person (3). Friday 28 of February 2020, the Dr. Manuel Martinez Baez Institute of Epidemiological Diagnosis and Reference (InDRE) confirmed the first case of COVID-19 in Mexico. They found others four cases with suspicious symptoms in people whose had been traveled to Italy, of which three of them had mild symptoms. Following this up, two of these patients are citizen of Mexico City and the last one is resident of Sinaloa State. The fourth patient did not show coronavirus symptoms; he was reported as a virus carrier. Perhaps this was the first Mexican case registered of an asymptomatic patient (3).In the next days, there were confirmed new cases of the pandemic. In Saturday 29 of February 2020, it was reported a female patient who tested positive while she was returned from Italy and she lives in Torreon, Coahuila Mexico. Her symptoms were mild and she stayed at home for her cares. The next case was shows up in Tuxtla Gutierrez, Chiapas, it was an 18 years old female patient, who also arrived from Italy and she was been in contact with the Torreon’s patient case. Until March 1st, all the COVID-19 Mexican cases were imported (3).

## METHOD

### COVID-19 Data Analysis

This present section has an analysis of the COVID-19 risk from Mexico’s regions. For the record, it is important to point out that the database origin is from the official government web page created by the Secretary of Health in Mexico ^4^.

The Mexico COVID-19 database has the following hierarchical variables (see Figure 1): 1) positive patients and negative patients, 2) symptomatic patients, 3) hospitalized patients and non-hospitalized patients.

**Figure 1:**
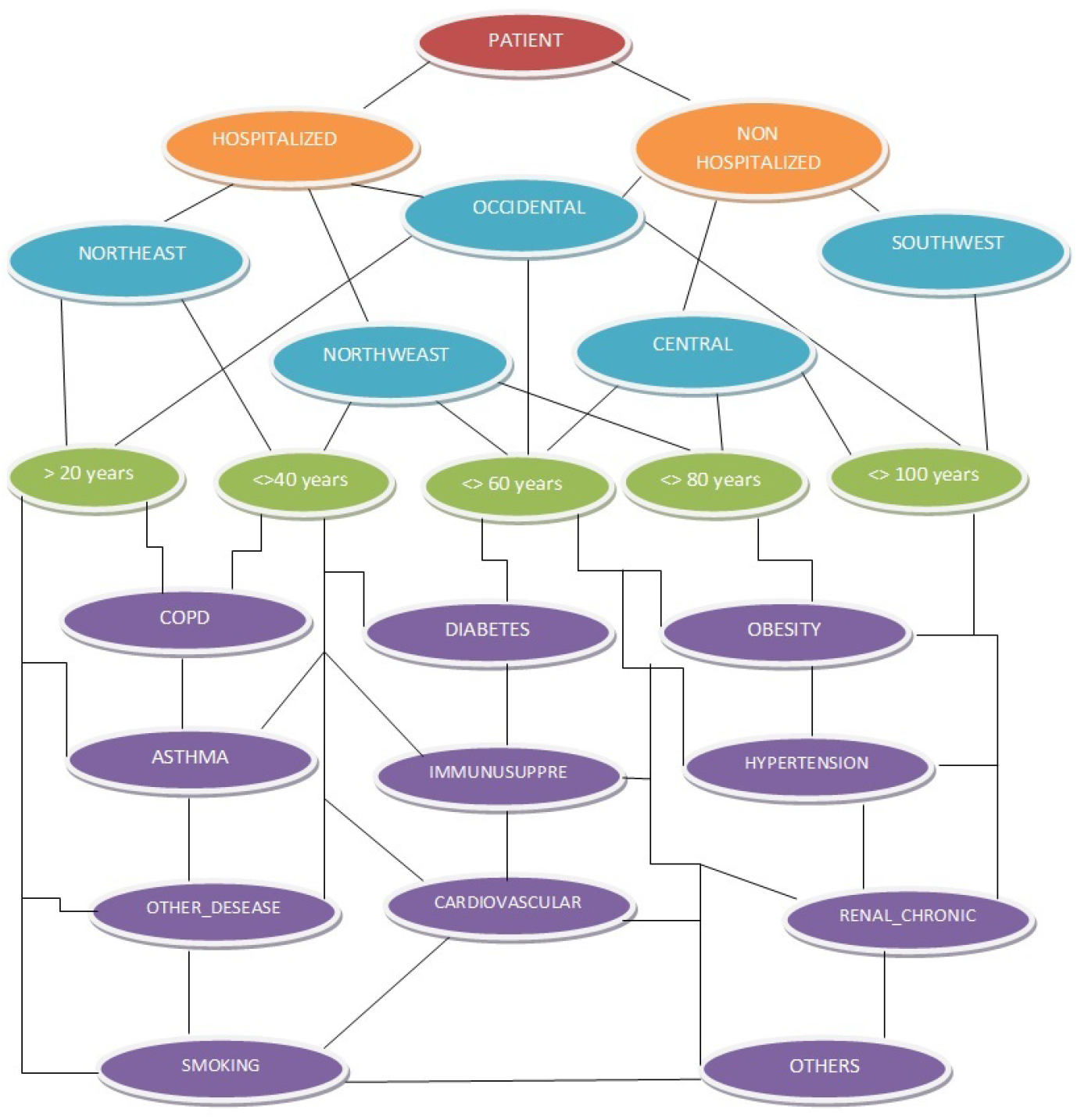
Network database were constructed hierarchical variables identified by 5 different colors as follows: maroon, orange, blue, green and purple.

Symptomatic patients are characterized of present the major COVID-19 symptoms, these patient cases have developed such as cough, sore throat, fever or shortness of breath. Once the viral detection test has been applied, if the patients are tested positive they were classified as positive patient and assume to have the virus, in other hand they are negative patients. At the first position on the hierarchical variables are positive patients; in these cases are subsections as follows: the symptom onset date, clinic admission date and clinic exit date (5). At the second place on the hierarchical variables are symptomatic patients whose are subsections as hospitalized patients and non-hospitalized patients. For symptomatic patients with severe to critical illness or who are severely immunocompromised the health experts ingress them at the clinic immediately and they are classified as hospitalized patients in our database. For symptomatic patients with mild to moderate illness who are not severely immunocompromised the health experts recommended that they must keep a strict quarantine at home and they are classified as non-hospitalized patients in our database. The study project report is based in a compared of the hospitalized and non-hospitalized patients with the comorbidities of the patients and their exposure risk to the virus in different Mexico’s regions. In these statistical analysis, the principal comorbidities on patients are detailed, such as diabetes (D), COPD (CO), Asthma(A), Immunosuppression (IM), Hypertension (HY), cardiovascular (CA) problems (5), chronic kidney disease (RE), obesity (OB), among others diseases (OD) (6, 7). People who suffering of any comorbidities are at increased risk of severe illness from COVID-19, the diseases mentioned above play an important role in the possible recovery of patients who have acquired the virus (8). Figure 1 describes the Mexico COVID-19 database extraction and their hierarchical variables (9). It should be emphasized that the period of analysis of the database correspond from April 12 to June 29, 2020 (220, 667 patients), giving a total of 79 daily records files.

## STATISTICAL ANALYSIS

### Mexico’s Regions Parameters Analysis

For the Mexico’s regions risk analysis on Patients with COVID-19 were selected 5 regions, for a more adequate analysis and information management (11, 12). It must not forget that Mexico has 32 states, as a whole are each sovereign jurisdictions, each ones with a particular political division, economic entity, specific culture and population also with different social situation. The states grouped by separate regions allow an efficient and certain descriptive analysis of statistical information. Being on the same page, the regions are named as successive: United Mexican States of Northwest Region (R1), United Mexican States of Northeast Region (R2), United Mexican States of West Region (R3), United Mexican States of Central Region (R4) and United Mexican States of Southeast Region (R5).

The United Mexican States of Northwest Region (R1) consists of the next federal entities: Baja California, Baja California Sur, Chihuahua, Sinaloa and Sonora. The United Mexican States of Northeast Region (R2) incorporates: Coahuila, Durango, Nuevo Leon, San Luis Potosi and Tamaulipas. The United Mexican States of the western Region (R3) involves: Aguascalientes, Colima, Guanajuato, Jalisco, Michoacán, Nayarit, Querétaro and Zacatecas. The United Mexican States of the Central Region (R4) contains: Mexico City, State of Mexico, Guerrero, Hidalgo, Morelos, Puebla and Tlaxcala. Finally, the United Mexican States of Southeast Region (R5) includes the federal states of Campeche, Chiapas, Oaxaca, Quintana Roo, Tabasco, Veracruz and Yucatán.

### Mexico’s Regions Age Analysis

Giving support to the Mexico’s regions age analysis section, as in the previous partition division, now in this section, it was divide the database of age in five ranges (bins) were each bin corresponds to the next categories: 0−20 years old, 21 − 40 years old, 41 − 60 years old, 61 − 80 years old and 1 − 100+ years old (see Figure 2). The age distributions analysis compares the hospitalized (H) and non-hospitalized (NH) patients in the 5 Mexico’s regions where they were registered daily (13). The age distribution has been adjusted to obtain a normal distribution (13, 14). The first analysis contains the mean and standard deviation per day and then these analyses were calculated per groups for the consequent 79 days in the five geographical regions. The Figure 2 shows only 10 days of the 79, for more details visit our repository on GitHub.

**Figure 2:**
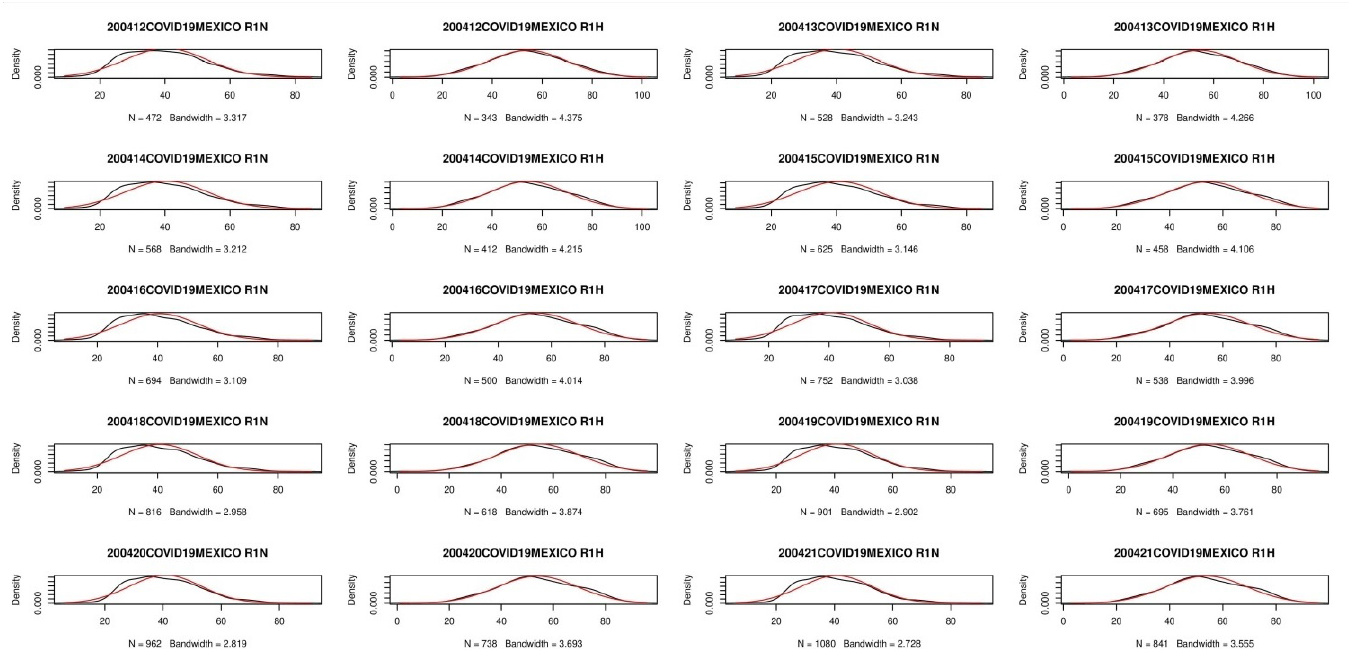
Age distributions plot of H and NH patients in R1 for 10 days.

It is important to mention that the next numbers are referring to the standard deviation age from the patients. In R1 the age of non hospitalized patients is about 40.88 years (standard deviation), while in the case of hospitalized patients it is almost 55.25 years old. In R2, NH patients have almost 40.64 and 54.64 years old for hospitalized patients. For R3 NH patients the age is 41.08 and for hospitalized patients is 55.46 years old. In R4 NH patient’s age is 42.42 and H patients are about 53.71 years old. For R5 the age of non-hospitalized patients is 42.22 and for hospitalized patients is 54.18 years old. It is notable in the age distribution that two bins accumulate the major age’s density and they are in 20–40 years old range and 40-60 age ranges. For the bin with range ages between 20 to 40 years, our analysis gets as result with the major density of NH COVID-19 patient cases, where the health experts recommend that they must keep a strict quarantine at home for their recovery. In contrast, for the bin with range ages between 40 to 60 years, our analysis gets as result with the major density of H patient cases. The Figure 2 carries the empirical evidence of these results, it is extraordinary that each region has independence between them, and it is amazing to discover that age average numerical and its age standard deviation value are similar in all regions.

### Mexico’s Regions Comorbidity Analysis

Giving backing to Mexico’s regions comorbidity and its analysis section, it is necessary to notice that these analysis were carry out only in H patients due to the huge information that can be able to obtain of them in the hospitals even each patients have been monitored all day. As we mentioned in the previous subsection, the age of these patients is between 40-60 years. The total number of patient registered in R1 are about 28,769 in these 79 days (*NH* = 18, 526, *H* = 10, 243), in R2 there were 20,178 patients (*NH* = 16, 752, *H* = 3, 426), for R3 with almost 26,359 (*NH* = 19, 544, *H* = 6, 815), R4 with 105,744 patients (*NH* = 69, 807, *H* = 35, 937) and R5 were 39,617 patients (*NH* = 27, 737, *H* = 11, 880). Chronic degenerative diseases have been a risk factor on patients with COVID-19. Depending on the comorbidity patient condition it is his scenario when the coronavirus show up, with fatal scenarios the risk is too high even they can die. The comorbidity scenarios registered by the Secretary of Health in Mexico are: patients with asthma, cardiovascular problems, COPD, diabetes mellitus, hypertension, immune problems, obesity, kidney problems and others. Figure 3 has two graphs with the Box plot where were compared the 5 Mexico’s regions and their morbidities and the patients are divided in hospitalized and non-hospitalized (14). In a critical observation there are highlight at least three most frequent degenerative comorbidities stand out among H and NH patients: obesity, diabetes mellitus and hypertension. It should be noted that in the R4 has more density and variety of comorbidities in cases of patients than in the R2. In the next analysis it has been presented the Relative Frequency of the Total number of comorbidities patients with COVID-19 for each ones Mexico’s Regions with the following Formula: Region = {Non-hospitalized, Hospitalized}.

**Figure 3:**
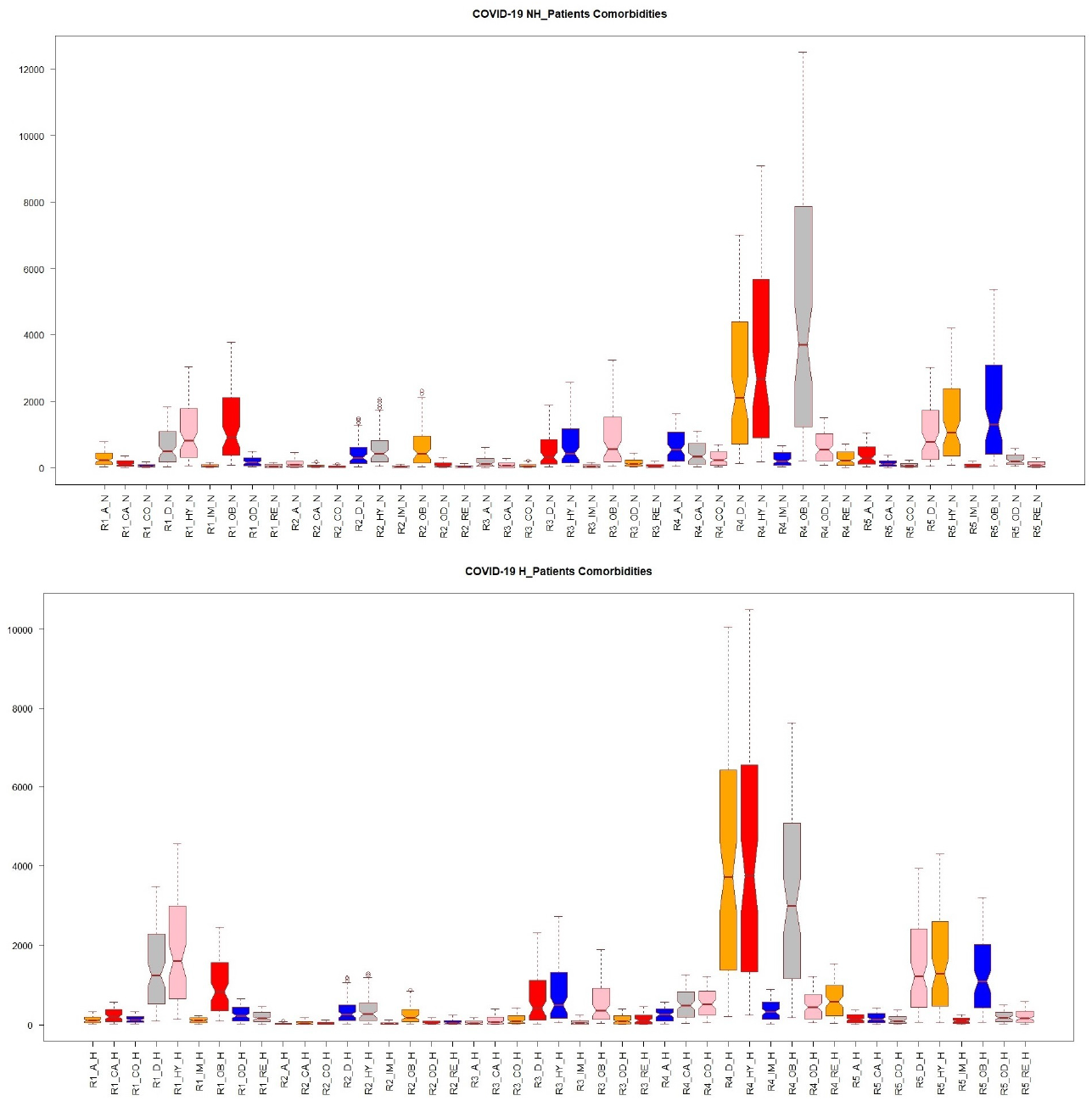
Box plot of the 5 Mexico’s regions grouped by the 9 comorbidities, the top graph is from NH patients values and the bottom graph is form H patients.

Asthma disease has had a relative frequency in patients with COVID-19 in Mexico’s regions of: *R*1 = {4.18%, 3.16%}, *R*2 = {2.72%, 2.65%}, *R*3 = {3.12%, 2.71%}, *R*4 = {2.31%, 1.60%} and *R*5 = {3.82%, 3.11%}. Cardiovascular diseases disease has had a relative frequency in patients with COVID-19 in Mexico’s regions of: *R*1 = {1.92%, 5.62%}, *R*2 = {1.07%, 5.42%}, *R*3 = {1.45%, 5.82%}, *R*4 = {1.59%, 3.52%} and *R*5 = {1.33%, 3.62%}. Hypertension diseases disease has had a relative frequency in patients with COVID-19 in Mexico’s regions of: *R*1 = {16.35%, 44.56%}, *R*2 = {12.26%, 37.53%}, *R*3 = {13.12%, 40.04%}, *R*4 = {13.01%, 29.17%} and *R*5 = {15.14%, 36.17%}. COPD disease has had a relative frequency in patients with COVID-19 in Mexico’s regions of: *R*1 = {0.93%, 3.13%}, *R*2 = {0.68%, 3.53%}, *R*3 = {1.11%, 6.17%}, *R*4 = {0.99%, 3.37%} and *R*5 = {0.81%, 3.11%}. Diabetes Mellitus disease has had a relative frequency in patients with COVID-19 in Mexico’s regions of: *R*1 = {9.83%, 33.95%}, *R*2 = {8.85%, 34.55%}, *R*3 = {9.65%, 33.82%}, *R*4 = {10.04%, 27.97%} and *R*5 = {10.87%, 33.21%}. Immunosuppression disease has had a relative frequency in patients with COVID-19 in Mexico’s regions of: *R*1 = {0.86%, 2.21%}, *R*2 = {0.71%, 3.38%}, *R*3 = {0.86%, 3.40%}, *R*4 = {0.93%, 2.47%} and *R*5 = {0.68%, 2.13%}. Obesity disease has had a relative frequency in patients with COVID-19 in Mexico’s regions of: *R*1 = {20.34%, 23.93%}, *R*2 = {13.81%, 25.27%}, *R*3 = {16.56%, 27.82%}, *R*4 = {17.90%, 21.27%} and *R*5 = {19.32%, 26.91%}. Kidney disease has had a relative frequency in patients with COVID-19 in Mexico’s regions of: *R*1 = {0.92%, 4.45%}, *R*2 = {0.82%, 7.03%}, *R*3 = {1.01%, 6.76%}, *R*4 = {1.02%, 4.25%} and *R*5 = {1.07%, 4.86%}. Others disease has had a relative frequency in patients with COVID-19 in Mexico’s regions of: *R*1 = {2.63%, 6.45%}, *R*2 = {1.90%, 5.34%}, *R*3 = {2.23%, 5.91%}, *R*4 = {2.16%, 3.38%} and *R*5 = {2.07%, 4.15%}.

It is worth noting that the major comorbidities densities that are presented in patients with COVID-19 are obesity, diabetes mellitus and hypertension diseases. The Table I is a resume of the 5 Mexico’s region patients; it shows the percentage patients for each region whose have been positive for COVID-19 but they are not having any type of the comorbidities. Of these patients, the 61%-69% is NH patients while 29%-41% required hospitalization it means, they are H patients.

**Table 1:**
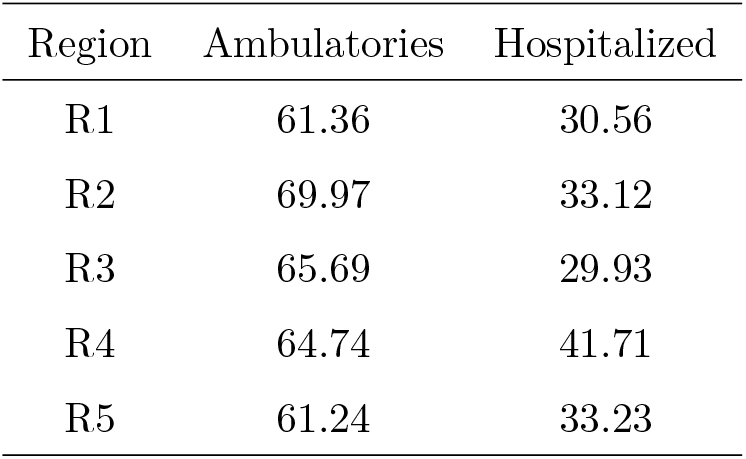
It contains the records of the patients for each Mexico regions whose did not report having any comorbidity when they tested positive to the virus.

| Region | Ambulatories | Hospitalized |
| --- | --- | --- |
| R1 | 61.36 | 30.56 |
| R2 | 69.97 | 33.12 |
| R3 | 65.69 | 29.93 |
| R4 | 64.74 | 41.71 |
| R5 | 61.24 | 33.23 |

### Mexico’s Regions Covariances Analysis

This subsection gives out an analysis of the dependencies of comorbidities with the different regions and their patients’ ages. The correlation network is constructed by hierarchical variables that are grouping: at the first level is the patient that tested positive for COVID-19; at second level it is divided into hospitalized and non-hospitalized patients; in a third level are the five geographic Mexico’s regions; at fourth level there are five age ranges of the patients and finally, in the fifth level there are eleven comorbidities types (see Figure 1). The correlation matrix of these comorbidities was generated by Pearson’s correlation coefficients whose values are between in the interval *ρ ∈* [−1, 1] (16, 17). The correlation’s matrix obtained is a square matrix of 91 by 91 elements. For a better visual analysis of these matrices, it was divided into two sub-matrices (see Figure 4). Each sub-matrix speaks for the positive or negative correlation between those conditions (18).

**Figure 4:**
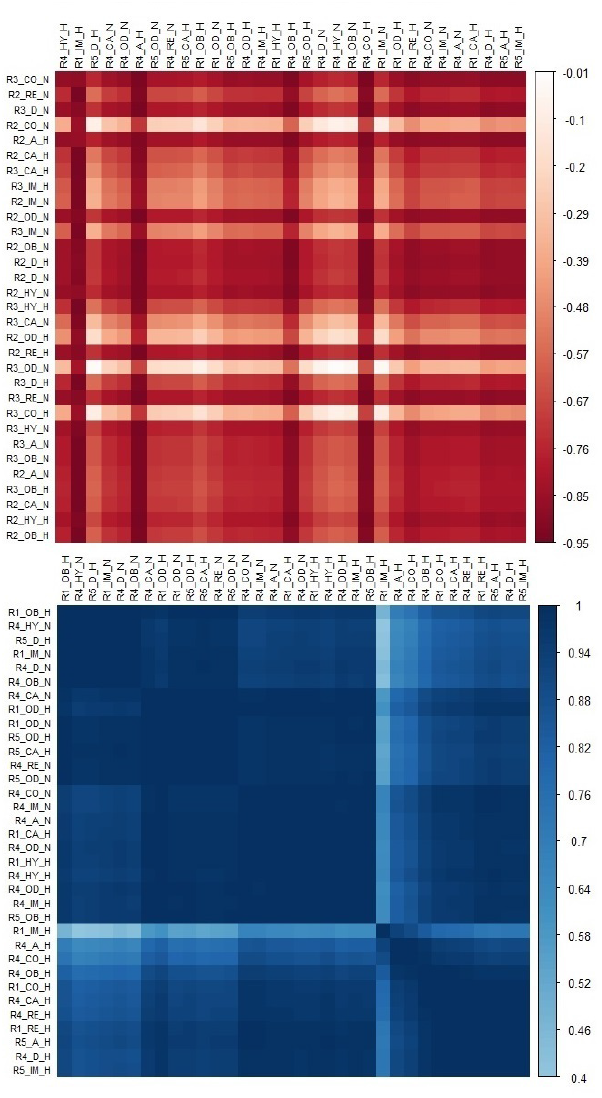
Correlation’s sub-matrices, the top matrix constitutes the positive correlation and the bottom matrix the negative correlation.

Figure 4 has 2 sub-matrices, the top matrix has contains the highest positives correlations that the analysis was found in the original matrix, the values almost are 1. In favor of last affirmation, it must be noted that his correlation are high because there are several hierarchical levels of the correlation network where the comorbidities are coincident and this has a positive effect on this relationship between H and NH patients. That is means, that the linear correlation is high due the patients in R5 with diabetes has similar density of H patients in R1 with obesity. In contrast, Figure 4 in the bottom has the matrix that represents the sub-matrix with the highest negative correlation that the analysis was found in the original matrix, that means, this sub-matrix has the inverse linear correlations between the hierarchical levels of the network. Figure 5 shows a correlogram map (19). These kinds of maps synthesize all the information analyzed in previous sections. Giving backing to last sentence, these correlogram has in its vertical axis the scale that represents the number of days (that is, all recorded database from the first day to day 79). On the below horizontal axis are the NH and H patients. The correlogram map contains the entire matrix correlation that has been described in previous sections, left vertical and upper horizontal axes describe the 5 hierarchical levels of correlation function in a kind of ramp palette. The correlogram’s color is a result of correlations patterns and they are determined by the density of the accumulated number of patients (20, 21).

**Figure 5:**
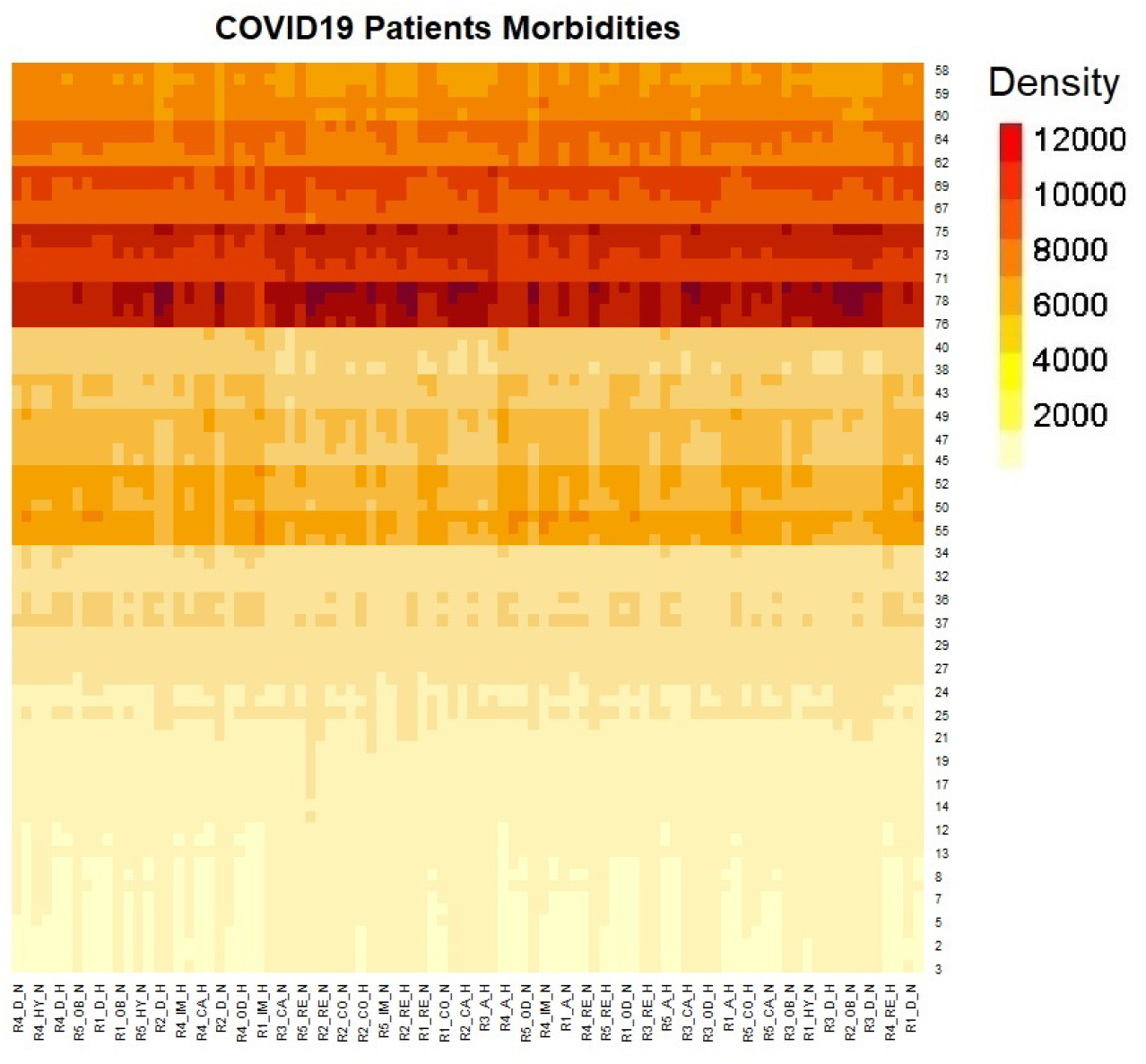
SuperHeat map describes the density relationship between the comorbidities, N and NH patients in the Regions of Mexico during the 79 days with the pandemic.

### Probability Analysis

This section proposes a method analysis of the probabilities that using the 9 subset comorbidities summatory and their intersections for each region of Mexico throughout the 79 days of analysis. The calculation of each probability is Equation 1, Equation 2 and Equation 3; this section also contains the description of the DAG plot and its relationship between the children nodes and its leaves, where were calculated the Bayesian probability Analysis. In the first case, it is important to note the direct relationship between the P (patients) and its type classification NH and H. Then the hierarchical classification is under 5 regions (R1, R2, R3, R4 and R5). Subsequently, the dependent event between the 9 comorbidities is calculated in two more hierarchical classifications (Comorbidities(H) and Comorbidities(NH)).

It is important to highlight that the depend evens are calculated by the probabilities of comorbidities and its intersections between each of them. It must be taken into account that the patients do not necessarily suffer only one comorbidity, in the majority of the situations they are suffering more than one comorbidity, see Figure 6. This phenomenon case is can be possible to resolve as it is presented in Figure 6. Note: There are two separate analysis, NH and H patients.

**Figure 6:**
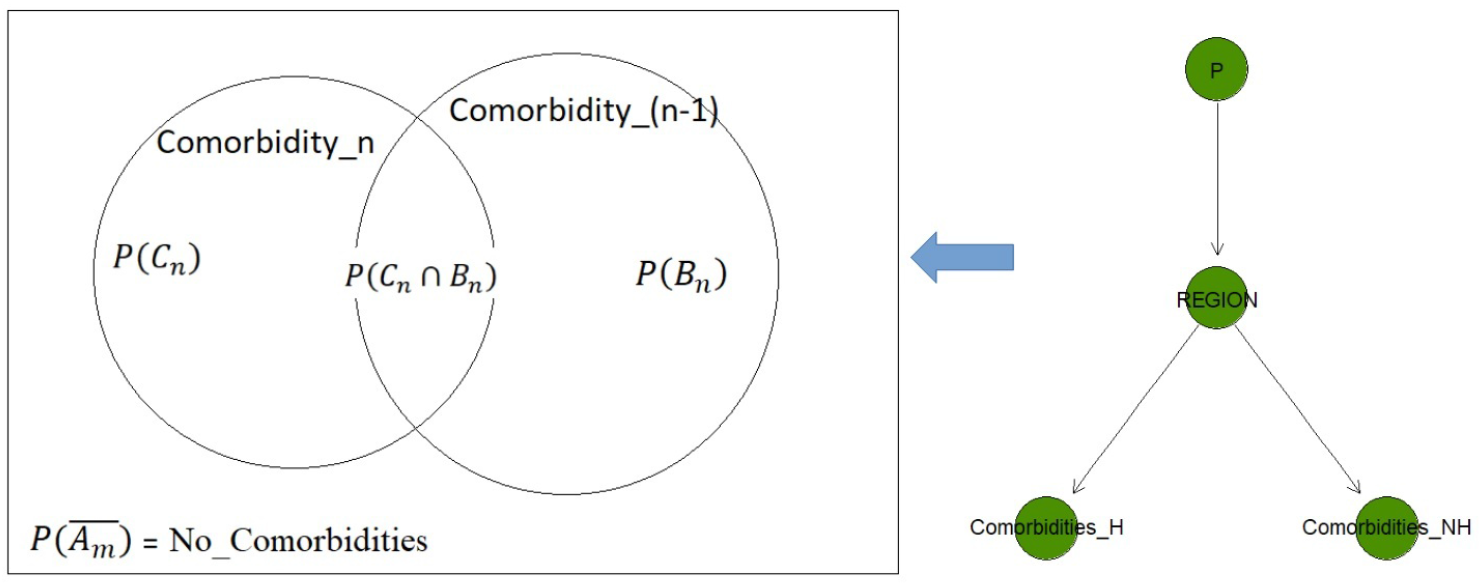
Bayesian Network configuration for calculate the Probability Analysis.

Defined to *P* (*A*_*m*_) as the probability of a patient with COVID-19 for the *m* = {1, …, 5} region. Therefore, 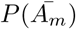 is defined as the complement probability, which is a patient in another region.

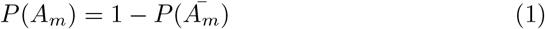

The intersection between regions and comorbidities is defined as:

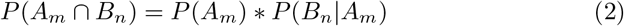

where *P* (*B*_*n*_|*A*_*m*_) is the region probability given at least one comorbidity *n* = {1, …, 9}. As consequence, the probability of each comorbidity is obtained by:

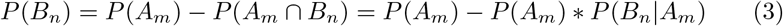

Based on the last equations (22**?**), these data results are entered in the Bayesian network that it uses the bnlearn and Rgraphviz libraries (in R programming) for generating the probabilities outputs that are shown in Figure 7 and Figure 8, for NH and H Patients.

**Figure 7:**
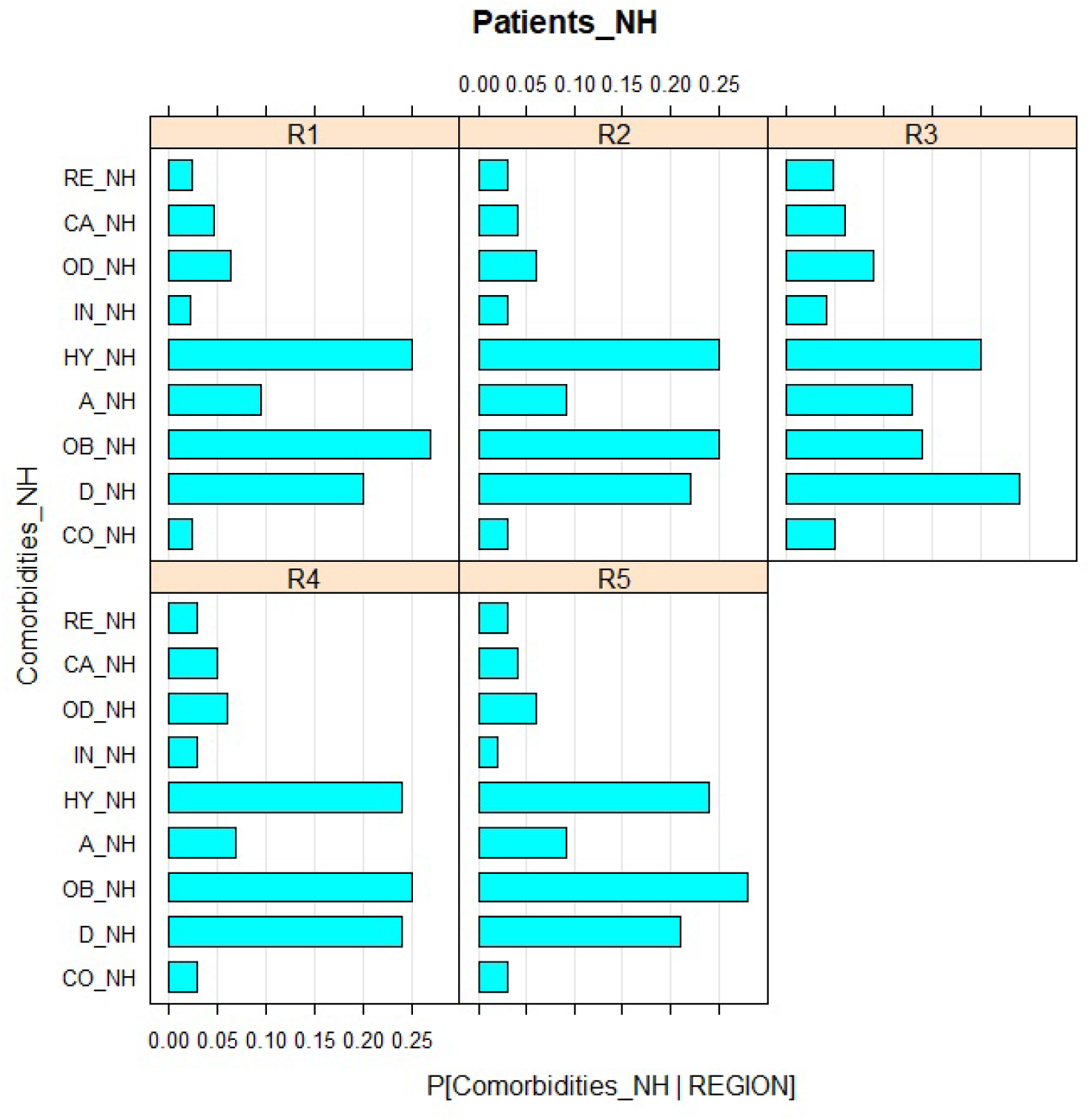
Network output for the Non-Hospitalized patients and their coefficients from comorbidity probabilities for the 5 Mexico’s regions in 79 days of COVID-19 registration.

**Figure 8:**
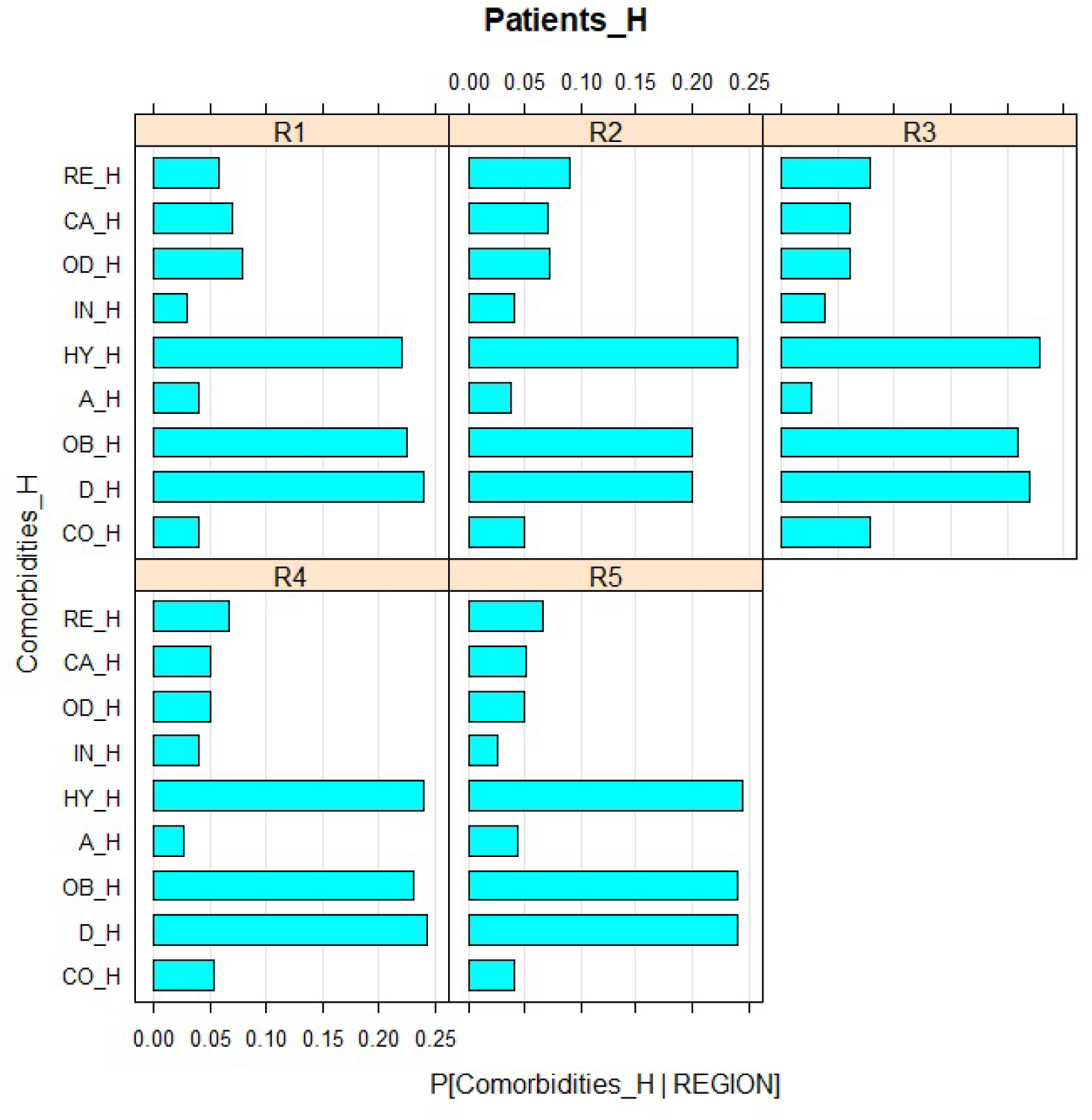
Network output for the Hospitalized patients and their coefficients from comorbidity probabilities for the 5 Mexico’s regions in 79 days of COVID-19 registration.

## DISCUSSION

In the last sections have described the study project report overview of the pandemic situation since the COVID-19 arrived to Mexico. The United Mexican States has a population of 125 million of people. Its largest concentration of the population is young and adult persons; as a consequence the elderly people are a lesser percent. Accordingly, there is a high probability that people who have been tested positive for COVID-19 are in the age range of 20-60 years old. In view of the fact that comorbidities occurs in adults people, the patients who have required hospitalization (H) are in the range about of 40-60 years old while patients who have not required hospitalization (NH) stay in the younger population 20-40 years old (see Figure 2).

In R1 35.61% of the patients have been hospitalized and 64.39% have not required hospitalized. For R2 16.98% is H and 83.02% NH; R3 the proportion of H is about 25.86% and NH with 74.14%; R4 have 33.99% of H while 74.14% are NH patients and in R5 it is about 29.99% of H patients and 70.01% of NH patients.

On the other hand, Mexican population have stood out in the comorbidities like as diabetes, COPD, asthma, Immunosuppression, hypertension, cardiovascular problems, chronic kidney disease, obesity, and others diseases. These illness conditions and COVID-19 have played an important role in determining whether or not patients require hospitalization. It is redeemable to mention that in R5, asthma, diabetes mellitus, kidney problems, and obesity have had a highest relative frequencies among H and NH patients. In R1 the cardiovascular problems, hypertension and others disease are the most frequent comorbidities in their patients, while in R4 the Immunosuppression is the mainly frequent comorbidity in their patients and COPD in R3. Some percent of H patients have been required mechanical ventilators in the Intensive Care Unit (ICU) in these 79 days. As a result, the accumulated relative frequency of invasive ventilator employed in H patients by region is as follows: 9.27% in R1, 10.47% in R2, 8.08% in R3, 7.45% in R4 and in R5 almost 10.79%. It should be emphasized that the patients whose needed the ICU services unfortunately died. The accumulated relative frequency in 79 days and their relationship is as follows: R1 of 48.42%, in R2 of 33.14%, R3 of 46.46%, for R4 of 56.49% and in R5 of 49.45%. Therefore, the risk of lose their life being H patients and ICU hospital services in R1 about 41.34%, R2 concerning 27.23%, 28.55% for R3, in R4 is 36.06% and R5 almost 35.31%. As claimed by the probability analysis, it can be possible to point out that there is a high probability that a good number of H patients are suffering obesity, hypertension or diabetes disease.

This research proposes a regional analysis of the risk of patients whose tested positive to COVID-19 and its relationship in a huge country like Mexico. This study also analyzed the risk exposure of patients with comorbidities in Mexican society by regional territory and it is notable the different risk values that involves the population. The evaluation period runs from April 12 to June 29, 2020 (almost 220,667 patients). The most densely populated R1 and R4 represent the largest number of positive cases that have required hospitalization. In R2 the major density of patients has recovered at home. As result of the age analysis distribution, the average age in NH patients is around 40 years old while in H patients it is close to 55 years old. While in R1 and R5 they have a higher proportion of H patients, in R2 have a superior proportion of NH patients. In other hand, H patients show that they are more sensitive due they have suffering comorbidities. And their previous scenario to arrived COVID-19 they have developed obesity, diabetes mellitus and hypertension and others disease. It is remarkable those H patients whose have needed a mechanical respirator have ranges about 7.45% to 10.79% for the all Mexico’s regions. Given that above scenario the risk of lose their life in the hospital is highest due to the aforementioned comorbidities, it is R1 and R4 being the region with the major propensity to die with COVID-19 patients while R2 is lower vulnerable than others, information obtained from the database statistical analysis. On the report of the analysis of probabilities (Figure 6) in all regions there is almost a 70% probability of being hospitalized if you have COVID-19 and suffer any type of the 9 comorbidities (Figure 7 and Figure 8), on the other hand there is a probability of almost 30% if you do not suffer any of the 9 comorbidities. In front of this national Mexican scenario, it is intended at work to generate a more detailed regional statistical analysis particularly in Mexico City (into R4), this state has the most densely populated territory of the country. This mentioned study allows to the Mexican population to take the necessary care to reduce the number of infections and therefore, the cases of deaths.

## Data Availability

All data is availability in a Github place.

https://github.com/OraliaNJ/COVID-19_Mex_Analysis

## Footnotes

4 Our file repository dataset inputs and their analysis are available on: https://github.com/OraliaNJ/COVID-19_Mex_Analysis

## Notes

### Competing Interest Statement

The authors have declared no competing interest.

### Funding Statement

None

